# Decoding pre-movement neural activity from thalamic LFPs for adaptive neurostimulation in tremor patients

**DOI:** 10.1101/2024.12.18.24319164

**Authors:** Fernando Rodriguez Plazas, Thomas G. Simpson, Laura Wehmeyer, Rahul S. Shah, Jamie Brannigan, Michael G. Hart, Pablo Andrade, Francesca Morgante, Veerle Visser-Vandewalle, Erlick A. Pereira, Huiling Tan, Shenghong He

**Affiliations:** Medical Research Council Brain Network Dynamics Unit, Nuffield Department of Clinical Neurosciences, University of Oxford, Oxford, UK; Neuromodulation and Motor Control Section, Neurosciences and Cell Biology Institute, City St George’s, University of London, London, UK; Nuffield Department of Surgical Sciences, University of Oxford, Oxford, OX3 9DU, UK; Department of Stereotactic and Functional Neurosurgery, Faculty of Medicine and University Hospital Cologne, University of Cologne, Cologne, 50937, Germany

**Keywords:** movement decoding, local field potentials, ventral intermediate nucleus, thalamus, tremor disorders, machine learning, deep brain stimulation

## Abstract

**Objective:** Understanding the neural mechanisms underlying movement initiation is crucial for advancing movement-driven adaptive deep brain stimulation therapies for tremor disorders. We investigated the feasibility of decoding pre-movement periods of upper limb movements by machine learning using thalamic local field potentials (LFPs) and scalp electroencephalography (EEG) signals.

**Approach:** Data were analysed from 11 patients undergoing deep brain stimulation surgery, employing machine learning models—including logistic regression, gradient-boosted decision trees, and convolutional neural networks—to distinguish rest periods from pre-movement periods.

**Main results:** Our results demonstrate that early neural correlates can predict movement onset, achieving above-chance decoding performance starting approximately 680 ms before movement execution using thalamic LFPs and 1.09 s using EEGs. Individualized, patient-specific decoders outperformed cross-patient models, reflecting substantial inter-patient variability in neural modulatory patterns. Additionally, multiple frequency bands contributed independently to decoding performance, emphasizing the importance of incorporating a broad spectrum of frequencies rather than relying solely on single canonical bands.

**Significance:** These findings underscore the value of personalized, multi-band machine learning approaches in capturing the neural correlates preceding movement. They support the development of adaptive neurostimulation therapies that enhance the effectiveness of clinical interventions through tailored models that account for patient-specific neural activity.

## 1. Introduction

Since receiving FDA approval for the treatment of essential tremor (ET) in 1997, deep brain stimulation (DBS) of the ventral intermediate nucleus (VIM) of the thalamus has established itself as an effective therapy for the treatment of tremor disorders in patients with medication-refractory symptoms (Dallapiazza et al., 2019; Ferreira et al., 2019; Kremer et al., 2021). In a meta-analysis comprising 1,714 ET patients, VIM-DBS improved tremor scores by a mean 61.3% at 20 months follow-up, showing a sizeable therapeutic effect in terms of tremor suppression (Lu et al., 2020).

Despite its efficacy for tremor management, VIM-DBS is associated with several adverse effects, including stimulation-induced side-effects that often impact speech and postural stability, as well as a gradual loss of therapeutic efficacy over time (Alomar et al., 2017; Barbe et al., 2011; Becker et al., 2020; Fasano & Helmich, 2019; Pahwa et al., 2006; Paschen et al., 2019; Peters & Tisch, 2021; Petry-Schmelzer et al., 2021). To address these challenges, non-continuous or adaptive DBS (aDBS) schemes have been proposed as a means to mitigate stimulation-related side effects and thereby increase the therapeutic window (Cernera et al., 2021, 2024; Meidahl et al., 2017; Opri et al., 2020).

A critical aspect in the development of aDBS systems is the selection of an appropriate feedback signal to modulate stimulation in real time. Various signals have been explored for driving aDBS in ET patients, including muscle activity recorded from surface electromyography (Cernera et al., 2021; Graupe et al., 2010; Yamamoto et al., 2013), scalp electroencephalography (EEG) (Herron et al., 2015), and signals recorded from intracranial electrodes (Herron et al., 2017; Opri et al., 2020; Ferleger et al., 2020). However, there is growing interest in leveraging modulatory patterns found in neural signals recorded from the implanted DBS electrodes themselves to avoid the need for supplementary implants or external devices (Priori et al., 2013). By capturing the neural correlates of movement through local field potentials (LFPs) measured from the thalamus, it is possible to detect movement states and utilize this information to drive an aDBS system (Tan et al., 2019). This approach has been successfully implemented in an acute clinical setting in prior studies (Castaño-Candamil et al., 2020; Ferleger et al., 2020; He et al., 2021; Buijink et al., 2022).

In the context of aDBS for patients with intention tremors (a type of tremor that typically manifests during voluntary movement rather than at rest), the timing of stimulation initiation can be critically important. Early triggering of stimulation, specifically initiating stimulation prior to or at the very onset of voluntary movement, holds significant therapeutic potential. By delivering stimulation before the tremor becomes clinically apparent, it may be possible to prevent the tremor from emerging altogether. This pre-emptive approach contrasts with reactive paradigms that activate the stimulation in response to tremor after it has already manifested – which may inherently involve too long a delay to reduce symptoms. Early stimulation could modulate the neural circuits involved in tremor generation at a stage when they are more susceptible to intervention, potentially suppressing the pathological oscillations that give rise to tremor. By intercepting the neural processes leading to tremor, this strategy could reduce or eliminate tremor episodes before they begin, enhancing therapeutic outcomes. Achieving this, however, requires systems capable of decoding the onset of movement in real time by detecting neural correlates present in the period leading up to movement execution. Early detection of movement from neural signals has been demonstrated, with electrocorticography (ECoG) signals showing higher decoding performance than LFP signals recorded from the subthalamic nucleus of Parkinson’s disease (PD) patients (Köhler et al., 2024).

In this study, we aim to benchmark our ability to train machine learning models to decode early correlates of upper limb movement during the pre-movement period, using LFP signals recorded from thalamic DBS electrodes and scalp EEG electrodes. To this end, we develop and implement several classification algorithms, utilizing both manually extracted features and features derived through deep learning techniques. We compare the performance of these models and analyse their outputs to understand the neural correlates at the cohort level that inform their predictions.

The remainder of this manuscript is structured as follows: we first introduce the dataset, feature extraction pipeline, and classification algorithms used in our study. We then present the decoding performance achieved by these systems and ascertain the neural correlates that they rely on to make inferences. Finally, we discuss the trade-offs of the different approaches and their implications for early movement-driven aDBS in tremor disorders.

## 2. Methods

### 2.1. Participants and Experimental Protocol

Data were collected from 11 patients (5 female) diagnosed with movement disorders and scheduled for DBS surgery for tremor management. The cohort included nine patients with ET, one with Orthostatic Tremor (OT), and one with tremor-dominant PD. Clinical information for each participant is provided in Table 1. All procedures were conducted in accordance with the Declaration of Helsinki and approved by the relevant local ethics committees. Informed written consent was obtained from all participants prior to their inclusion in the study.

**Table 1:** Clinical details of patients.

| PATIENT ID | DIAGNOSIS | DBS SYSTEM | SEX | AGE [YEARS] | DISEASE DURATION [YEARS] | PREDOMINANT SYMPTOMS BEFORE SURGERY |
| --- | --- | --- | --- | --- | --- | --- |
| S1 | ET | Abb | F | 76-80 | 21 | Tremor, gait ataxia, tremor worse on right, upper limb and voice tremor |
| S2 | ET | Abb | M | 70-75 | 8 | Tremor, upper limb, with right worse than left, lower limb tremor |
| S3 | ET | Abb | F | 60-65 | 45 | Tremor, upper limb tremor left worse than right, voice tremor |
| S4 | ET | Abb | M | 70-75 | 5 | Tremor, upper limb left worse than right |
| S5 | ET | Med | F | 56-60 | 15 | Tremor in both hands, left hand worse than right |
| S6 | ET | Med | M | 70-75 | 10 | Tremor in both hands (stronger in right hand), some head tremor |
| S7 | ET | Med | M | 60-65 | 20 | Tremor |
| S8 | ET | Med | M | 70-75 | 16 | Tremor |
| S9 | ET | Med | F | 56-60 | 50+ | Tremor |
| S10 | PD | Med | M | 66-70 | 6 | Rigidity and tremor in right hand and arm |
| S11 | OT | Med | F | 60-65 | 9 | Orthostatic and action tremor in both hands |

Abbreviations: ET = Essential Tremor; PD = Parkinson's disease; OT = Orthostatic Tremor; SGH = St. George's Hospital; UHC = University Hospital Cologne; Abb = Abbott Infinity electrode; Med = Medtronic SenSight electrode.

Patients underwent stereotactic neurosurgery for bilateral implantation of DBS leads targeting the ventral VIM, zona incerta (ZI), or posterior subthalamic area (PSA) for the treatment of tremor. For research purposes, the DBS leads were temporarily externalized for up to seven days post-implantation, allowing for direct recording of neural signals before implantation of the implantable pulse generator (IPG).

Four patients (S1–S4) were implanted with the Abbott Infinity DBS system (Abbott Laboratories, US), which features segmented directional leads. Each lead contains eight contacts, with the two central levels comprising three directional segments each. During recordings, the three directional contacts at each level were fused to function as a single ring-mode contact to prevent amplifier saturation. This configuration resulted in four ring-mode contacts per hemisphere. LFPs were amplified and sampled in monopolar mode using a TMSi Porti amplifier (TMSi, The Netherlands), with a sampling rate of 2,048 Hz. Seven patients (S5–S11) were implanted with Medtronic SenSight directional leads (Medtronic Inc, US), also featuring segmented contacts at the two central levels. For these patients, all contacts, including individual directional segments, were recorded in monopolar mode, yielding eight contacts per hemisphere. LFP signals were acquired using a TMSi SAGA amplifier (TMSi, The Netherlands) at a sampling rate of 4,096 Hz. A reference electrode was attached to the wrist of each patient to serve as a common reference point for monopolar recordings across all modalities.

Scalp EEG recordings were obtained using electrodes placed at positions Cz, C3, C4, CPz, CP3, and CP4 (following the international 10–20 system), encompassing somatosensory, sensorimotor, and motor cortical areas. EEG signals were recorded synchronously with LFP signals. Surface electromyography (EMG) was recorded using bipolar electrodes placed over the forearm flexor and extensor muscles. EMG signals provided precise markers of upper limb movement onset, aiding in the temporal alignment of neural and behavioural data.

During recording sessions, patients performed a series of upper limb motor tasks designed to elicit voluntary movement and engage different muscle groups: rice pouring task (patients poured rice back and forth between two cups), pegboard insertion task (patients inserted pegs into a pegboard), and foam ball gripping task (patients tightly gripped a foam ball with one hand).

Each task was performed multiple times with rest intervals to prevent fatigue. Instructions were standardized, and patients were encouraged to perform movements at a comfortable pace. All signals—including thalamic LFPs, scalp EEGs, and surface EMGs—were recorded using custom-developed software tailored for high-fidelity electrophysiological data acquisition. The software synchronized data streams across modalities and annotated timestamps for task events. Recorded data were digitized and stored securely for offline processing and analysis, adhering to data protection and privacy regulations.

### 2.2. Data Pre-processing and Labelling

The EMG signals recorded from the forearm flexor and extensor muscles were utilized to determine the timing of movement initiation. For lateralized movements, such as the one-handed gripping task, EMG traces from the arm executing the movement were selected. For non-lateralized movements, including the pegboard insertion and rice pouring tasks, signals from both arms were used for analysis.

To enhance the detection of movement onset, the EMG signals underwent a series of pre-processing steps. Raw EMG signals were first high-pass filtered using a causal fourth-order Butterworth filter implemented in second-order sections. To minimize ringing artifacts due to filter warm-up, the initial filter state was adjusted appropriately. All filtering was performed in a forward-only (causal) manner to preserve the temporal integrity of the signals. After high-pass filtering, the magnitude of the EMG signals was computed to obtain the envelope of muscle activity, and z-scored to standardize the amplitude across trials and participants. These processed EMG traces were visually inspected to ensure clear delineation of movement execution, and traces that did not distinctly define and isolate movement onset were discarded to maintain data quality. For tasks involving both arms, the EMG signals from each arm were averaged to create a composite signal representing overall muscle activity. This signal was then smoothed, and a threshold was set at the earliest points at which a deviation from baseline activity was detected (i.e. movement initiation). These timings were subsequently reviewed manually to ensure movement was detected at the earliest possible time point. This manual labelling served as the ground truth for subsequent analyses.

To prevent the introduction of unwanted phase lags between the movement signals and the neural signals, the same high-pass filter applied to the EMG data was also applied to the thalamic LFP and cortical EEG signals. This approach ensured temporal alignment across different kinds of signals. Additionally, to eliminate power line interference and its harmonics, notch filters were applied at 50 Hz, 100 Hz, and 150 Hz using a causal filter design with second-order sections.

The movement onset times defined from the EMG signals were used to segment the LFP and EEG data into epochs corresponding to individual movement trials. For participants who performed multiple movement-related tasks (gripping, rice pouring, and pegboard insertion), the epochs from all tasks were combined for analysis.

### 2.3. Time-Resolved Characterization of Oscillatory Activity

To train neural decoders capable of detecting pre-movement periods, we implemented a feature extraction pipeline (Figure 1) designed to capture relevant information within electrophysiological signals in a time-resolved manner. The extracted features aim to provide a comprehensive characterization of the relevant oscillatory activity at any given time point.

**Figure 1:**
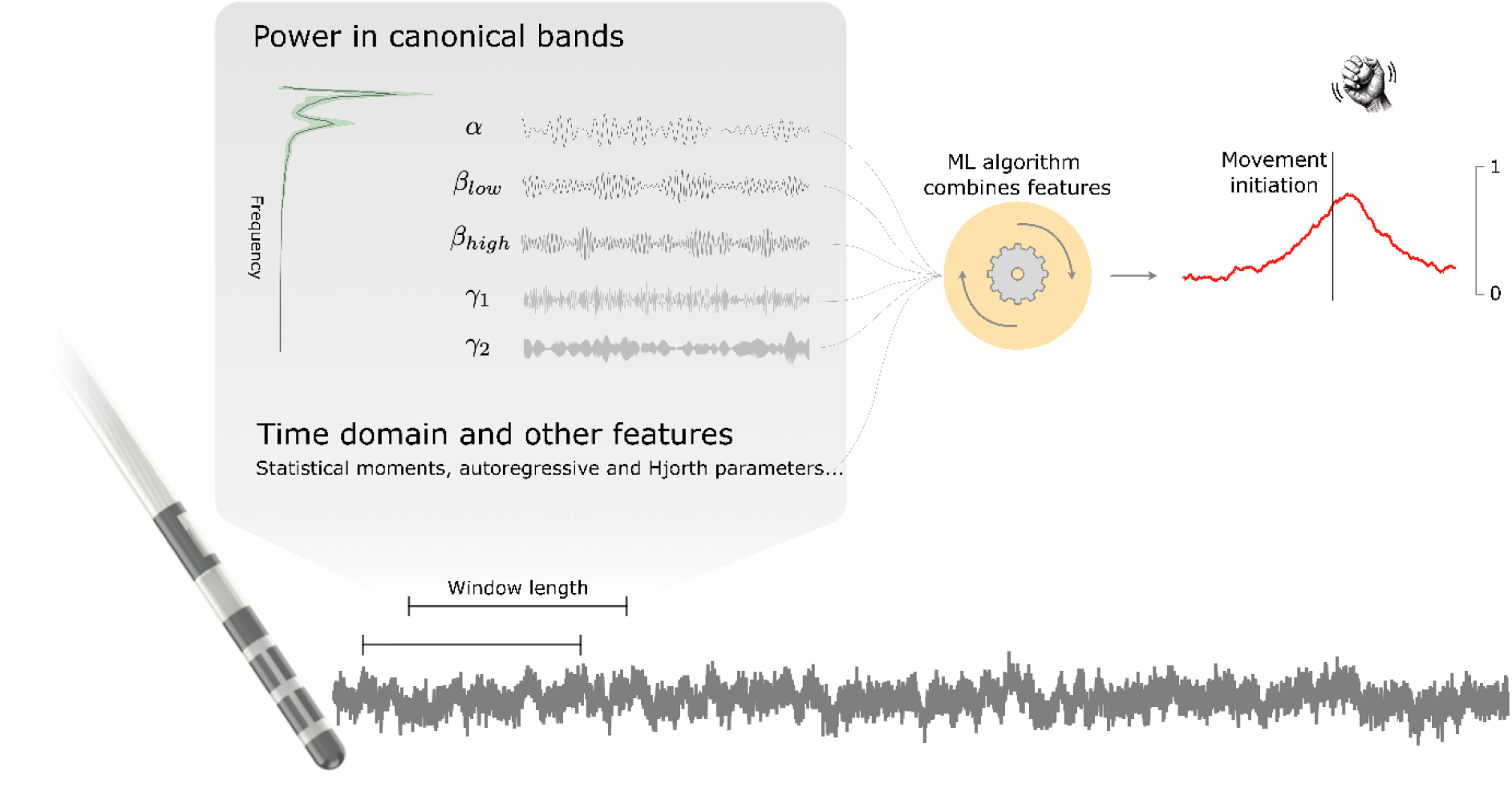
Illustration of the feature extraction pipeline used for decoding pre-movement periods from electrophysiological signals. Raw LFP and EEG signals are first pre-processed and segmented into overlapping trailing windows of 0.5 and 1-second lengths, updated every 20 milliseconds. Within each window, features are extracted, and the extracted features are normalized using causal running statistics and concatenated to form a feature vector. These feature vectors are used as inputs to machine learning classifiers designed to detect pre-movement periods in real time.

Specifically, time series signals were first segmented into short-term windows. From each of these windows, various features were extracted and subsequently concatenated along the time dimension, offering a time-resolved representation of the oscillatory activity contained within the signals. This overlapping windowing procedure is analogous to the short-term Fourier transform (STFT), a technique used to compute time-frequency decompositions. To avoid making strong assumptions about the optimal window length for capturing and representing the oscillatory dynamics of interest, we extracted features using two different window lengths: 0.5 seconds and 1 second. These features were provided to the classifier independently, granting the optimization algorithm access to features extracted over these two different time scales and allowing the classifier to weight them appropriately based on their contribution to detecting the state of interest.

To ensure that our analysis pipeline remained fully causal—that is, no information from future time points was used to make assessments at the present time point —the windows were computed in a trailing manner, i.e. utilizing only data points leading up to the current time point. We set the distance between contiguous windows at 20 milliseconds, which, in a real-time setting, corresponds to a system that outputs estimations at a rate of 50 Hz. This parameter allowed us to define the granularity of the state estimation pipeline, and a short distance between contiguous windows enabled us to capture and train our classifiers with the short-term fluctuations that may be encountered during real-time application.

At each time point, and for each trailing window length, we extracted several features to characterize the oscillatory activity within the electrophysiological signals comprehensively.

#### 2.3.1. Power in canonical frequency bands

For each time window of electrophysiological data, we computed a periodogram, which was subsequently log-transformed. Prior to conversion to the frequency domain, a Hann window was applied to the time-domain signals to minimize spectral leakage and mitigate edge artifacts that could distort the spectral estimates.

From the resulting periodogram, we extracted the mean and standard deviation of power within predefined canonical frequency bands. These bands were carefully selected to capture distinct oscillatory activities associated with various cognitive and physiological states. In our configuration, the bands included theta (4–8 Hz), alpha (8– 12 Hz), low beta (13–20 Hz), high beta (20–30 Hz), gamma (30–60 Hz, 60–80 Hz, 80–100 Hz), and high-frequency activity (200–500 Hz).

#### 2.3.2. Time-domain statistics

Time-domain statistics are features that capture the characteristics of the signal directly from its temporal representation. These features provide crucial insights into the underlying neural processes and are often used in the pre-processing or analysis stages of electrophysiological data. The primary time-domain features extracted included the **first four statistical moments** of the signal distribution as well as the Hjorth parameters.

The first four statistical moments are:

- **Mean (First Moment):** Average value of the signal over time. This provides a measure of the central tendency, the overall level of activity within the signal, as well as a measure of low-frequency activity.
- **Variance (Second Moment):** Computed as a Hjorth parameter (*Activity*, see below).
- **Skewness (Third Moment):** Quantifies the asymmetry of the signal’s amplitude distribution. A positive skew indicates a distribution with a longer tail on the right side, whereas a negative skew indicates a longer tail on the left. This measure detects deviations from normality in the signal’s amplitude.
- **Kurtosis (Fourth Moment):** Describes the “tailedness” or peakedness of the signal distribution. High kurtosis indicates the presence of infrequent, extreme deviations from the mean, often associated with transients or bursts in the signal.

The **Hjorth parameters** are a set of three descriptors useful in the analysis of non-stationary signals, such as LFP, EEG, or MEG data. They provide a compact representation of the signal’s dynamic properties:

- **Activity:** Corresponds to the variance of the signal, representing the overall signal power. It reflects the amplitude of the signal fluctuations and is directly related to the signal’s energy content.
- **Mobility:** Defined as the square root of the variance of the first derivative of the signal divided by the variance of the signal itself. It quantifies the mean frequency or the rate of change in the signal, with higher mobility indicating faster signal fluctuations.
- **Complexity:** Measures the signal’s waveform complexity relative to a pure sine wave, calculated as the ratio of the mobility of the first derivative of the signal to the mobility of the signal itself. It reflects the degree of variability in the frequency content of the signal, with higher values indicating more complex, non-sinusoidal waveforms.

Together, these time-domain features provided a comprehensive characterization of the electrophysiological signals, capturing essential aspects of the signal’s amplitude, variability, and temporal structure. They are commonly used in various applications, including brain-computer interfaces, cognitive state monitoring, and the identification of pathological activity such as epileptic seizures (Alawee et al., 2023; Hag et al., 2023; Hjorth, 1970).

#### 2.3.3. Other Features

We also extracted autoregressive (AR) parameters and cepstral coefficients to capture additional signal dynamics.

For the **autoregressive parameters**, we employed AR modelling to describe a signal as a linear combination of its 10 previous values. The AR parameters were estimated by solving the Yule-Walker equations, which relate the autocorrelation function of the signal to the AR coefficients.

The **cepstral coefficients** were derived through cepstral analysis, which involves taking the inverse Fourier transform of the logarithm of the signal’s spectrum, providing a representation in the *quefrency* domain. This technique can reveal periodic structures in the frequency domain. The cepstrum was segmented into 10 bands, and the power within these bands was computed, capturing the periodicity and harmonic structure of the signal.

Once all features were extracted, they were normalized by z-scoring using causal running statistics obtained from the previous 10 seconds of data. This normalization ensured that the features were on a comparable scale and that the analysis pipeline remained compatible with a real-time setting by avoiding introducing information from future time points (Merk et al., 2022).

### 2.4. Classification pipeline and evaluation of decoding performance

To train classifiers capable of detecting pre-movement periods, as shown in Figure 2, we labelled time windows ending within 500 milliseconds prior to movement initiation as belonging to the *pre-movement period*. Windows ending within the interval [−4, −2] seconds relative to movement onset were labelled as belonging to the *rest period*. Our classifiers were trained to distinguish between these two periods based on the features extracted from the corresponding electrophysiological time series. This paradigm focuses on detecting neural signatures preceding movement execution and allows for variability in the period of interest.

**Figure 2:**
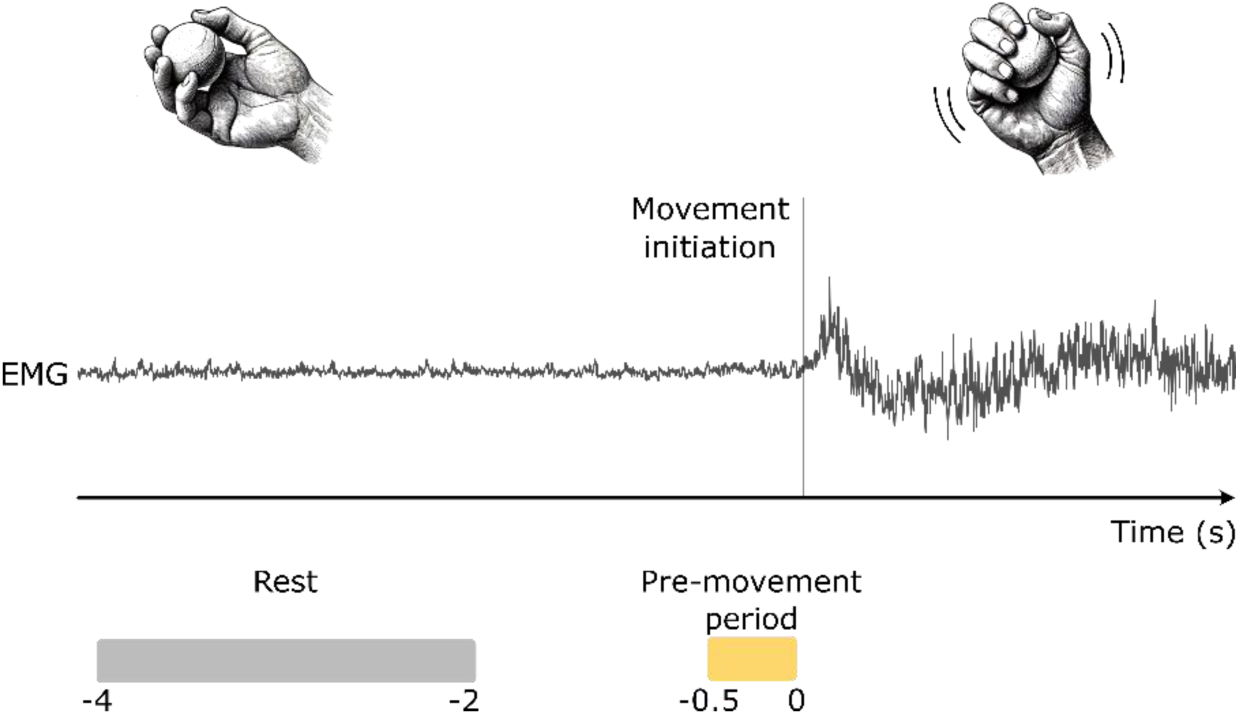
Illustration of the trial labelling process. Movement initiation is determined based on EMG activity. Feature extraction is strictly causal, ensuring that only features extracted from preceding samples are used for current estimations.

The performance of the classifiers was evaluated using a leave-one-trial-out cross-validation scheme. In each iteration, one trial was reserved as validation data to benchmark the classifier’s performance, while the remaining trials were used to train the classifier by setting its weights and parameters. This process was repeated iteratively, with each trial serving as the validation set once and as part of the training data in all other iterations. This approach ensures that all trials are used for out-of-sample testing, providing a robust estimate of the classifier’s generalization performance.

During each iteration, we computed the area under the Receiver Operating Characteristic curve (ROC-AUC; abbreviated AUC) for the predictions made by the trained classifier on the validation data. This metric was then averaged across all folds of the cross-validation to obtain a single cross-validated performance score. To account for class imbalances—specifically, the longer duration of the rest period compared to the pre-movement period— we adjusted the ROC curve by differentially weighting samples based on the period from which they were extracted. This weighting corrects for the disproportionate number of samples from the rest period. This procedure was performed individually for each EEG/LFP channel.

We then implemented and benchmarked four following classification architectures: logistic regression, gradient-boosted decision trees, convolutional neural network, and convolutional neural network with manually extracted features.

#### 2.4.1. Logistic Regression

Logistic Regression (LR) is a simple yet powerful classification algorithm that models the probability of a binary outcome using a linear combination of input features. The logistic regression model computes a linear combination of the features and applies the logistic (sigmoid) function to produce an estimate of the probability that the input features belong to the positive class *p*(*y* = 1 | *features*).

In our implementation, during each iteration of the leave-one-trial-out cross-validation, we performed an internal three-fold cross-validation on the training data to determine the optimal level of regularization (e.g., L1 or L2 penalty terms). During training, sample weights were adjusted to correct for class imbalance by giving more weight to samples from the minority class. Using the selected hyperparameters, a final logistic regression model was trained on the training data and evaluated on the validation trial for the current fold.

#### 2.4.2. Gradient-boosted Decision Trees

Gradient-Boosted Decision Trees (GBDT) is an ensemble learning method that builds a predictive model by sequentially combining multiple weak learners—typically decision trees—with each new tree aiming to correct the errors of the preceding ensemble. This iterative process is guided by gradient descent optimization, minimizing a specified loss function such as the negative log-likelihood.

In our implementation, we tuned the GBDT model to find the optimal hyperparameters, including the number of trees, learning rate, and tree depth, through an internal cross-validation procedure at each iteration of the leave-one-trial-out scheme. Specifically, we utilized a grid search to explore a range of hyperparameter values, selecting the combination that yielded the best cross-validated performance on the training data. Once the optimal hyperparameters were determined, the GBDT model was trained on the entire training set of the current fold and evaluated on the validation trial. Sample weighting was applied during training to account for dataset imbalance.

Our primary motivation for benchmarking GBDTs alongside logistic regression was to assess the potential benefits of capturing non-linear relationships between features and labels. If significant non-linear relationships existed in the data, GBDTs were expected to better capture and leverage these patterns compared to the linear logistic regression model.

#### 2.4.3. Convolutional Neural Network

Convolutional neural networks (CNN) are deep learning models that automatically learn hierarchical feature representations from raw input data through multiple layers of convolutional filters. Unlike the feature-based methods described above, CNNs take minimally pre-processed electrophysiological time series as input, without relying on manually extracted features.

In our CNN architecture (Figure 3), the input time series were processed through a cascade of convolutional layers interleaved with non-linear activation functions, batch normalization layers (to correct for scaling differences and improve training stability), and pooling layers (to reduce the dimensionality of activations). The convolutional filters were trained to extract relevant information content from the signals pertinent to the classification task. The training process involved the use of adaptive moment estimation (Adam), an adaptive gradient descent algorithm that iteratively adjusted the filter coefficients to minimize the cross-entropy loss between the model’s predictions and the true labels.

**Figure 3:**
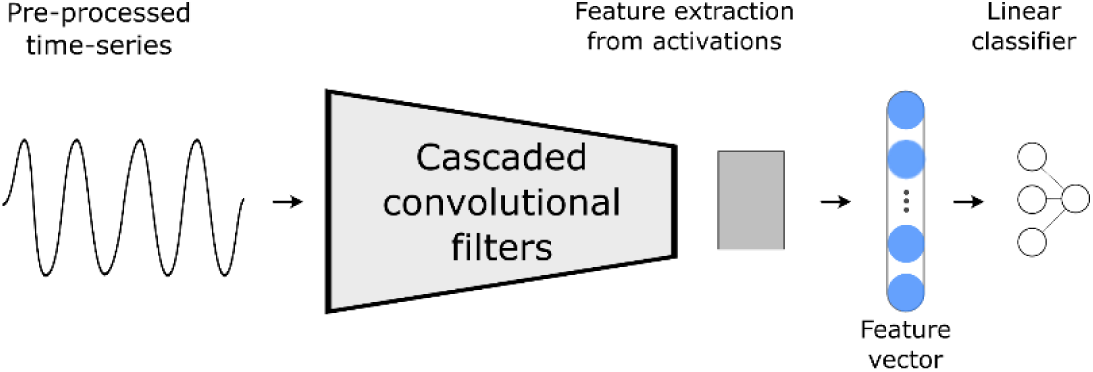
CNN Architecture. Minimally pre-processed electrophysiological time series signals are processed through a cascade of convolutional filters. Activations are filtered, scaled, combined, and passed through non-linear activation functions as they progress through the layers of the network. Final activations are summarized using descriptive statistics, which serve as input features for a linear classifier, producing the final logit.

At each stage of the network, the processed signals (referred to as “activations”) were filtered, scaled, combined, and passed through non-linear activation functions. After the final convolutional layer, descriptive statistics were computed from the activations, producing scalar features that were input into a fully connected linear classifier (perceptron), yielding the final output logits.

#### 2.4.4. Convolutional Neural Network with manually extracted features (FeatCNN)

To leverage both automatically learned features and manually extracted features, we implemented a hybrid architecture that combines the CNN described above with the manually extracted features from our feature extraction pipeline. After the time series data were processed by the convolutional part of the network, the resulting convolutional features were concatenated with the manually extracted features. This combined feature set was then fed into the fully connected linear layers at the end of the network (Figure 4).

**Figure 4:**
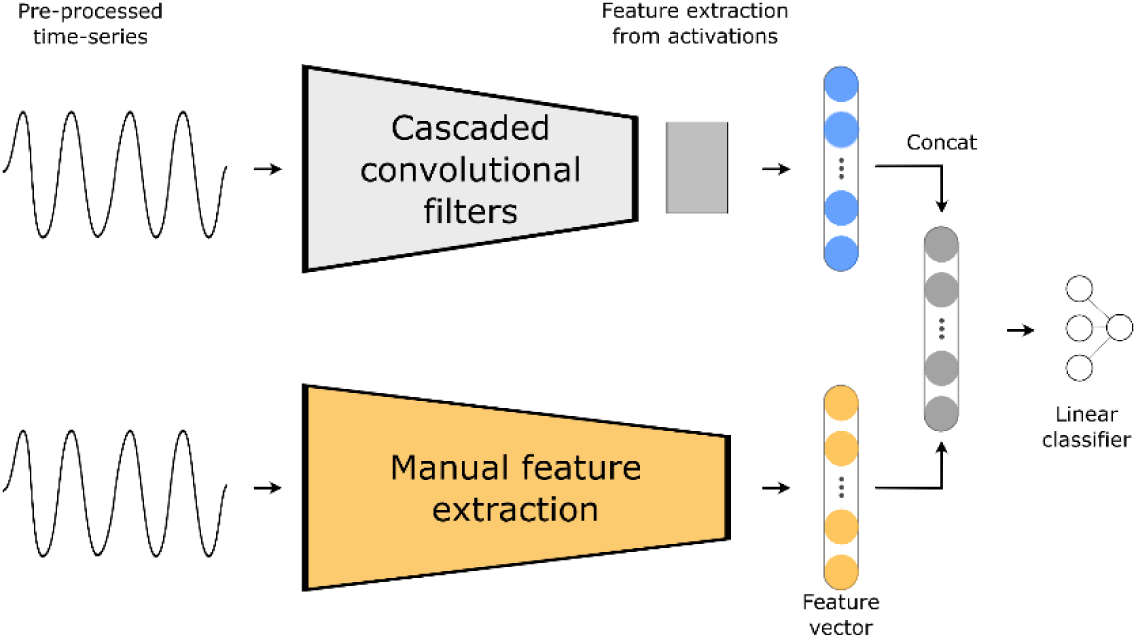
Feature-CNN Architecture, which combines a CNN and manually extracted features into a single model.

This approach allowed us to compare the performance of a model driven solely by learned convolutional features (CNN) with a model that integrates both manually extracted features and learned features (FeatCNN). By evaluating both architectures, we aimed to assess whether the addition of manually extracted features could enhance classification performance.

For both convolutional network architectures, the electrophysiological time series signals were down-sampled to 512 Hz from the original sampling rates (2048 Hz or 4096 Hz) prior to being passed to the network. The cascade of convolutional filters consisted of 6 layers, each with a convolutional filter comprising 45 kernel elements and a stride of 2 samples, a batch normalization layer, and a sigmoid linear unit (SiLU) activation function (Ramachandran et al., 2017). During training, we employed data augmentation techniques to enhance model generalization. Specifically, we performed data augmentation in the logit space by linearly combining samples using a manifold mix-up technique (Verma et al., 2019). This approach regularizes the model by encouraging smoother decision boundaries.

To address dataset imbalance during training, we applied a weighted sampling technique for the two CNN-based algorithms. Samples from the minority class (pre-movement period) were oversampled to ensure that the classifier received a balanced representation of both classes during training iterations.

Further details on the motivation, implementation, and suitability of these convolutional neural network models can be found in our prior work (Rodriguez et al., 2023).

## 3. Results

### 3.1. Decoding of Pre-Movement Periods

We evaluated the performance of four classification models—LR, GBDT, CNN, and FeatCNN—in distinguishing rest periods from pre-movement periods using both thalamic LFPs and scalp EEG signals.

Figure 5A illustrates the area under the Receiver Operating Characteristic curve (AUC) values for each model using LFP and EEG data. Higher AUC values indicate better decoding performance. Mean AUC values and the corresponding 95% confidence intervals are presented in Table 2.

**Table 2:** Mean AUC value [95% CI] for decoding the [-500ms, 0] pre-movement period with different models. Higher values indicate better decoding performance.

|  | LR | GBDT | CNN | FeatCNN |
| --- | --- | --- | --- | --- |
| LFP | 0.64 [0.60, 0.67] | **0.68 [0.65, 0.70]** | 0.64 [0.60, 0.68] | 0.65 [0.61, 0.68] |
| EEG | 0.62 [0.59, 0.65] | **0.68 [0.65, 0.71]** | 0.65 [0.61, 0.68] | 0.64 [0.62, 0.67] |

Paired *t*-tests were conducted to compare the performance of the models, with corrections for multiple comparisons applied using the False Discovery Rate (FDR) method. For the LFP-driven models, GBDTs significantly outperformed the other models. Specifically, the comparison between LR and GBDT (*p* = 0.002), between CNN and GBDT (*p* = 0.035), and between FeatCNN and GBDT (*p* = 0.033) indicated significant differences. No significant differences were found among the other three models (LR, CNN, FeatCNN).

For the EEG-driven models, GBDTs also showed superior performance compared to LRs (*p* = 0.0003). However, the differences between GBDT and CNN (*p* = 0.13) and between GBDT and FeatCNN (*p* = 0.03) were not statistically significant after correcting for multiple comparisons. Here again, no significant differences were observed among the other three models.

To determine the earliest time point at which movement onset could be predicted, we trained logistic regression models using a sliding 500 ms window, shifted in 25 ms increments from –2.25 seconds up to 1.5 seconds relative to movement onset. This resulted in AUC values at each time step, providing a time-resolved measure of the model’s ability to distinguish pre-movement periods from rest, as shown in Figure 5B.

**Figure 5:**
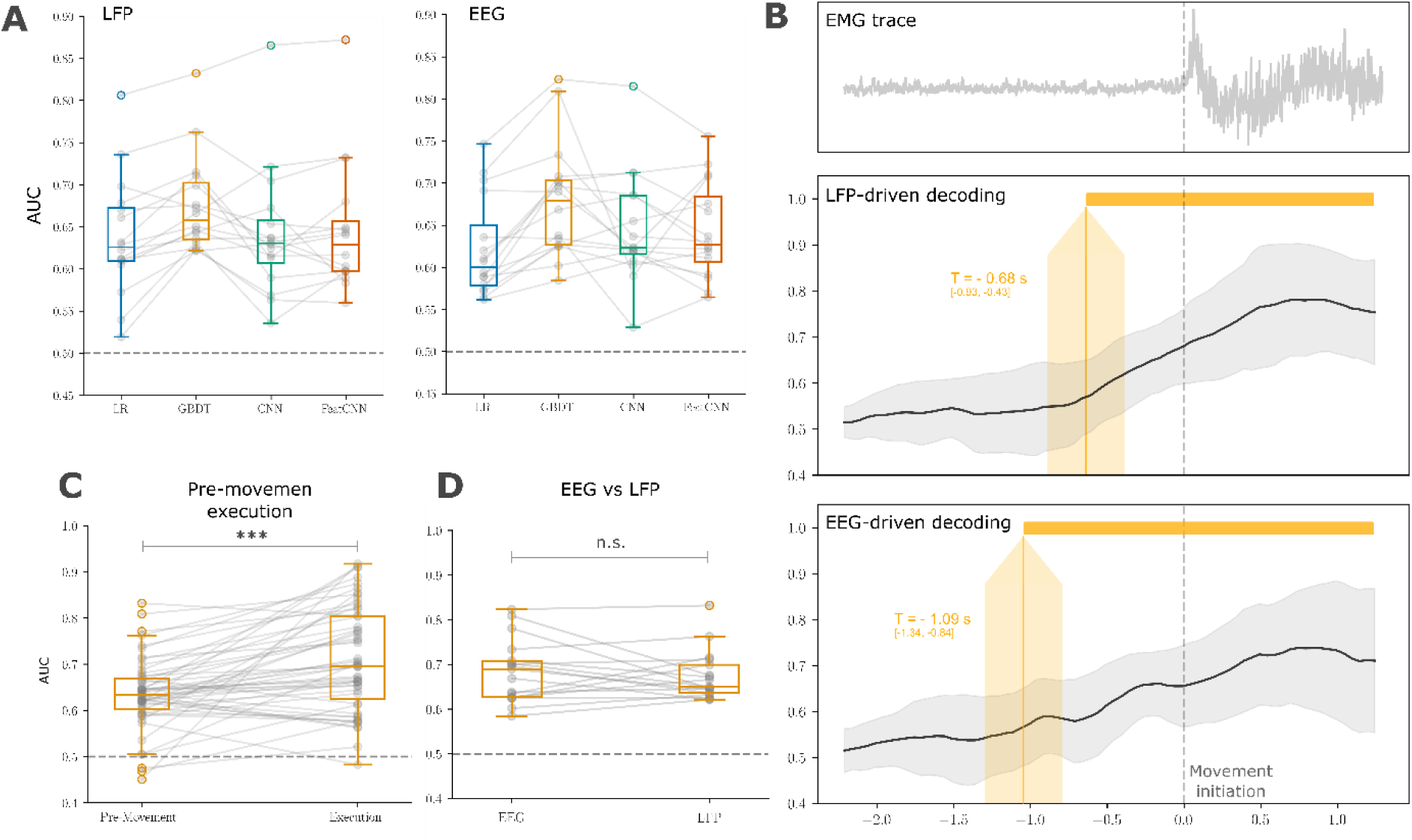
Decoding performance of different models in detecting pre-movement periods from neural data. (A) Classification model performance: boxplots of the area under the receiver operating characteristic curve (AUC) values for each classification model in distinguishing rest from pre-movement periods using LFP and EEG data. The boxplots represent the distribution of AUC values across participants. Individual dots outside the whiskers denote patients that are outliers. (B) Time-resolved decoding performance: Plots showing the time-resolved AUC values for logistic regression models trained to detect pre-movement periods at various time windows relative to movement onset. Movement onset (t = 0 s) is marked by the vertical dashed line. The top subplot displays the cross-participant average Electromyography (EMG) trace, aligning the timing of muscle activation. The middle (LFP data) and bottom (EEG data) subplots illustrate the progression of mean AUC values over time. The solid black lines represent the mean AUC across participants, and the grey shaded areas indicate ±1 standard deviation. The vertical yellow line and shaded region highlight the earliest time point at which decoding performance becomes significantly above chance level (AUC = 0.5), demonstrating each model’s ability to predict movement onset in advance. (C) Pre-movement vs. movement execution decoding: Comparison of AUC values for GBDT models trained to detect pre-movement periods (from –500 ms to 0 ms relative to movement onset) versus movement execution periods (0 ms to 500 ms) using LFP data. This comparison assesses the model’s effectiveness in decoding neural activity immediately before movement initiation versus during movement execution. (D) Comparison between signal modalities: AUC values for GBDT models utilizing LFP and EEG data, illustrating the difference in decoding performance between the two neural signal modalities. The results highlight the relative effectiveness of invasive (LFP) versus non-invasive (EEG) recordings in predicting pre-movement neural states.

Statistical analyses revealed that, for LFP-driven models, the first time window where the AUC was statistically significant above chance (paired *t*-test, *p* < 0.05) was the [–0.93 s, –0.43 s] interval, centred at 680 ms before movement onset. For EEG-driven models, the earliest significant window was [–1.32 s, –0.84 s], centred at 1,090 ms before movement onset.

We compared the decoding performance of GBDT models trained to detect pre-movement periods ([–500 ms, 0]) with those trained to detect movement execution periods ([0, 500 ms]) using LFP data. As shown in Figure 5C, for LFP-driven models, the AUC values for movement execution were significantly higher than for pre-movement decoding. A paired *t*-test yielded a *t*-statistic of 14.12 and a *p*-value less than 0.001, with an average AUC increase of 15.3% when comparing movement execution to pre-movement decoding. For EEG-driven models, the AUC difference was also significant, with a *t*-statistic of 9.02 and a *p*-value below 0.001, and an average relative AUC increase of 14.5%.

We assessed whether there was a consistent difference in decoding performance when using LFP signals from the thalamus versus EEG signals recorded from the scalp. Paired *t*-tests revealed no significant differences in AUC values between LFP and EEG across all models. The uncorrected *p*-values were 0.55 for LR, 0.47 for GBDT, 0.80 for CNN, and 0.87 for FeatCNN. The AUC values for GBDT models using LFP and EEG data are depicted in Figure 5D.

### 3.2. Cross-Patient Variability in Modulatory Patterns

Regardless of their architecture, classification models leverage changes in the information content of electrophysiological signals—often manifested as synchronization or desynchronization of activity in canonical frequency bands—to drive decoding performance. To visualize the average modulatory patterns across patients, we computed cohort-level time-frequency decompositions (see Figure 6). These were calculated using resonator IIR filters with a quality (Q) factor of 20, rectified, and smoothed using a 250ms kernel. Baseline corrections were applied using values extracted from the rest period defined as [–4, –2] seconds relative to movement onset. Additionally, we computed the average time-frequency decompositions separately for the top and bottom 50th percentile performers based on decoding accuracy to explore potential differences in modulatory patterns associated with classification performance.

**Figure 6:**
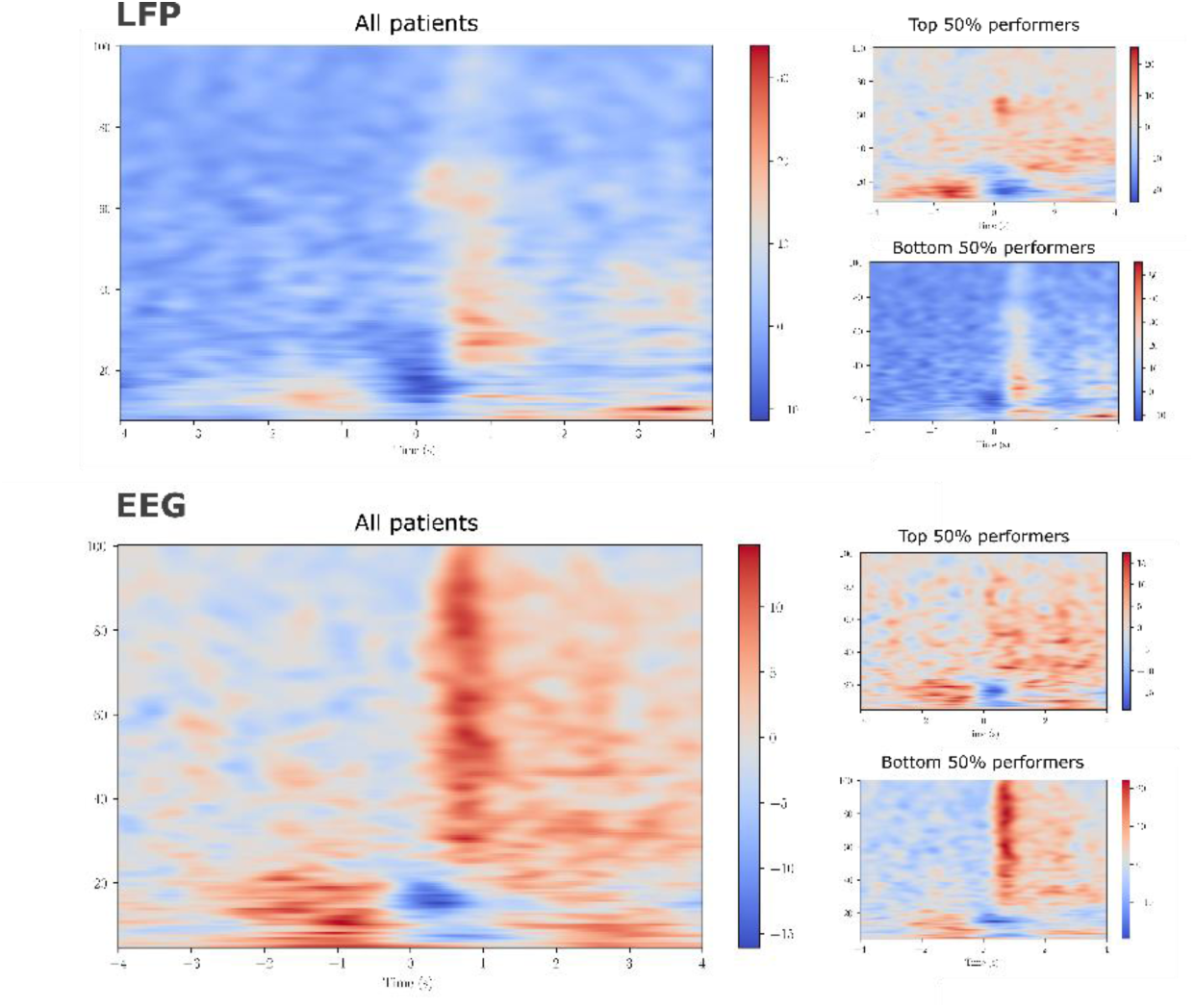
Time-frequency representations of cohort-level oscillatory activity around the onset of movement.

To investigate the specific modulatory patterns associated with the pre-movement period, we computed modulation indices for individual frequency bands. The modulation index for a specific band was calculated by comparing the power spectral density (PSD) estimates in that band during the pre-movement period with those during the rest period, averaged across trials:

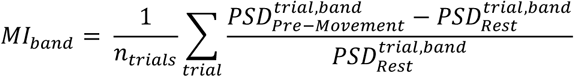

Modulation indices for each subject and band, along with the mean and 95% confidence intervals, are depicted in Figure 7A for both LFP and EEG signals. A negative modulation index corresponds to desynchronization (decreased activity) in the respective band during the pre-movement period compared to rest, while a positive modulation index indicates synchronization (increased activity). Figure 7B shows time-frequency decompositions from individual patients, illustrating the variability in modulatory patterns observed across the cohort.

**Figure 7:**
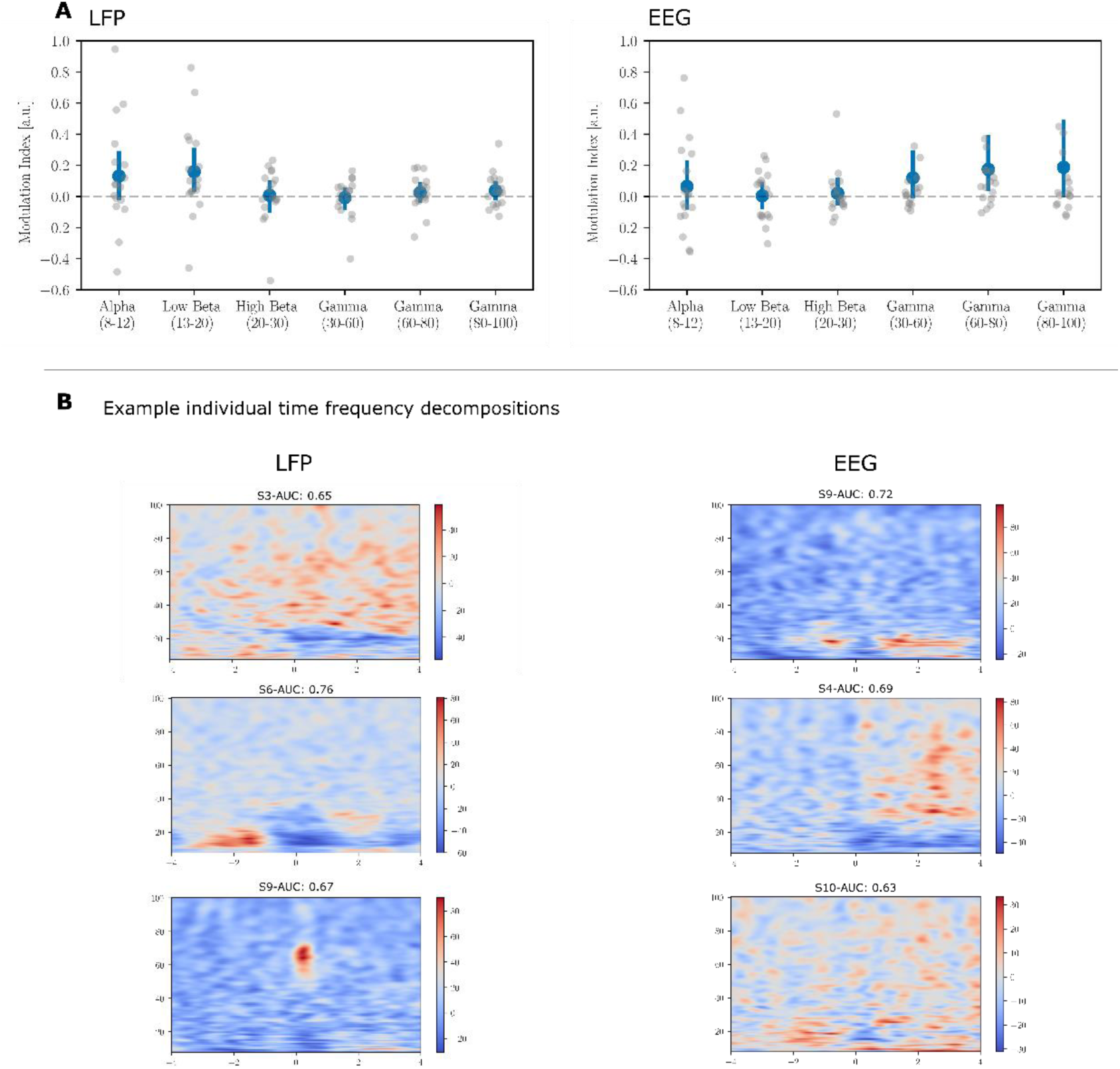
Cross-patient variability in the observed modulatory patterns. (A) Modulation indices for each band. Each dot represents an individual patient, and the blue bars represent the cross-patient mean and 95% confidence intervals. Negative modulation indices represent desynchronization (decreased activity) during the pre-movement periods, whereas positive indices represent synchronization (increased activity) relative to rest. (B) Individual time-frequency decomposition for a selection of patients, showcasing various modulatory patterns that are present in the dataset.

To assess the generalizability of modulatory patterns across patients, we trained logistic regression models under different training conditions and compared their out-of-sample performance. Specifically, we evaluated three training scenarios:

1. **Patient-Specific Training**: Models were trained exclusively on data from the patient of interest.
2. **Combined Training**: Models were trained on data from the patient of interest supplemented with data from other patients.
3. **Cross-Patient Training**: Models were trained exclusively on data from other patients, without including any data from the patient of interest.

We then evaluated the performance of these models on the patient-specific test data. For both LFP and EEG signals, including data from other patients during training led to a degradation in decoding performance compared to patient-specific training (*p* < 0.001 in both cases; see Figure 8). Furthermore, models trained exclusively on data from other patients performed significantly worse than those trained on combined data (LFP: *p* = 0.011; EEG: *p* = 0.0017). These results suggest that modulatory patterns associated with pre-movement periods exhibit substantial inter-patient variability, limiting the generalizability of models across patients.

**Figure 8:**
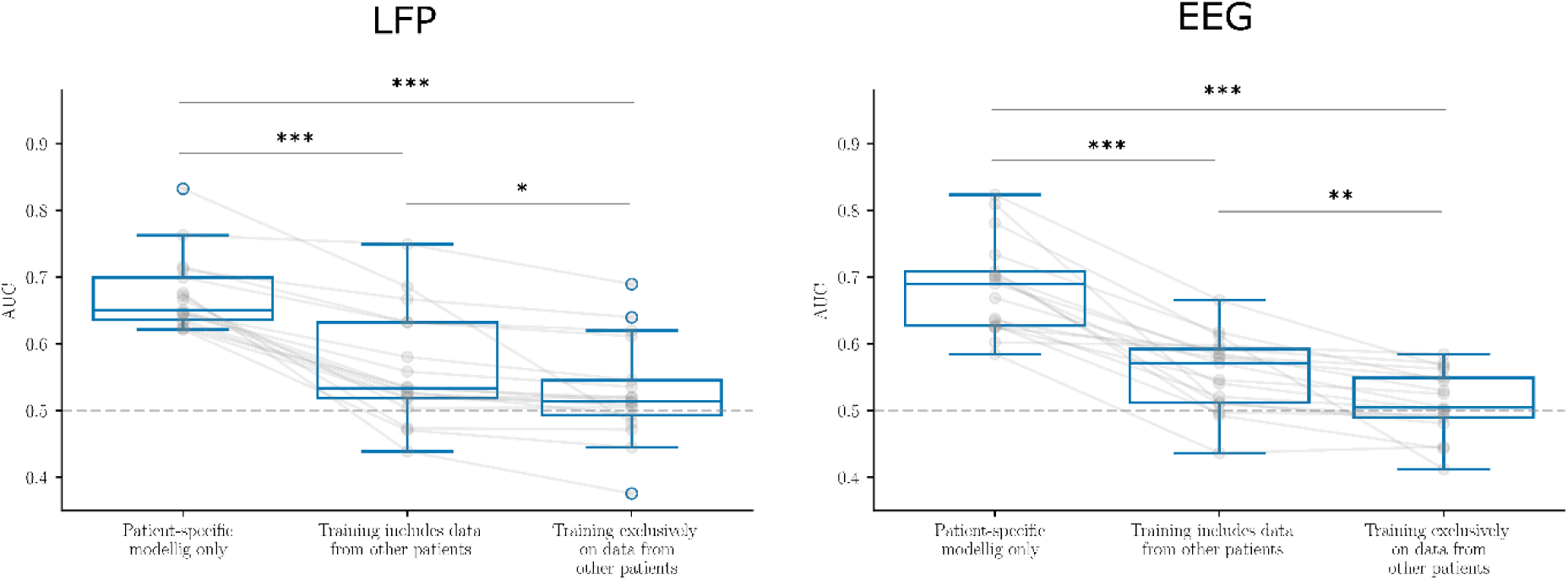
Logistic Regression models were evaluated on patient-specific test data, but trained under three different conditions: (1) using only patient-specific training data, (2) using combined training data that included both the patient’s data as well as data from other patients, and (3) using only training data from other patients. Decoding performance decreased when data from other patients was included during training, indicating limited generalizability of modulatory patterns across patients.

## 4. Discussion

### 4.1. Decoding Performance is Driven by a Broad Spectrum of Frequencies

In this study, we investigated the relationship between modulations in canonical frequency bands and the ability of our trained systems to detect pre-movement periods. We employed modulation indices to quantify changes in PSD within specific frequency bands during pre-movement periods compared to rest. We then attempted to correlate these modulation indices with the decoding performance, measured by the AUC values of LR models.

Figure 9 displays linear regressions between AUC values of LR models and corresponding modulation indices in canonical bands for both LFP and EEG signals. None of these regressions yielded *p*-values below the significance threshold of 0.05 (no correction for multiple comparisons was applied here). This lack of significant relationships suggests that no single frequency band consistently drives decoding performance across patients. Instead, contributions to decoding appear to be distributed across several frequency bands and may vary among individuals.

**Figure 9:**
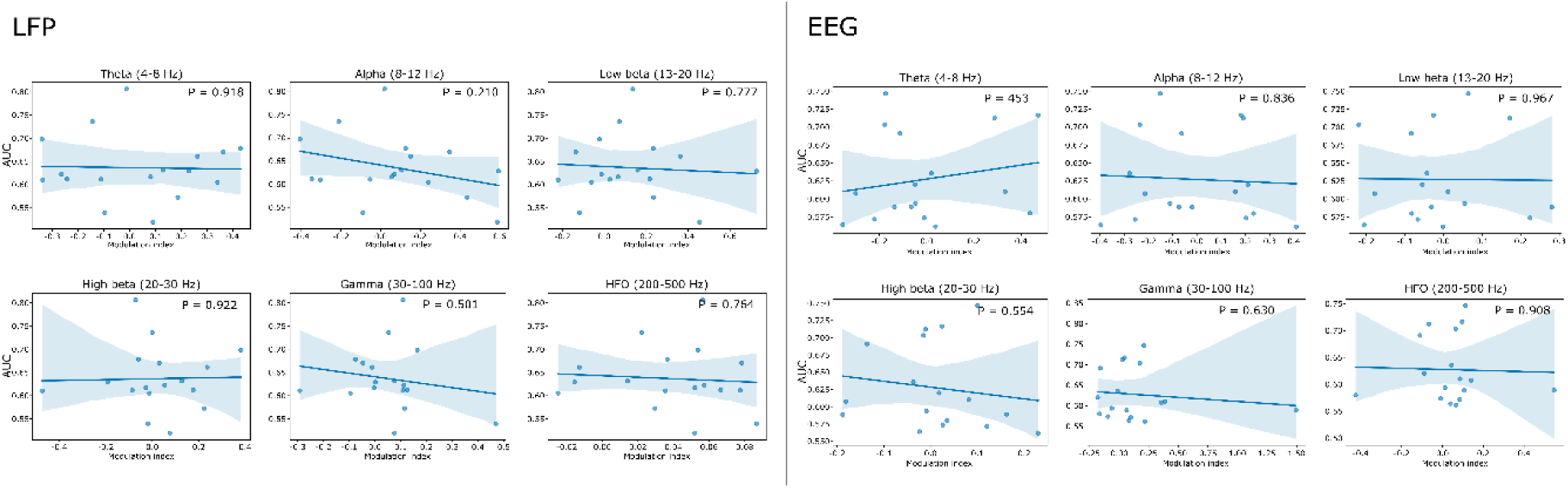
Correlations between modulation indices and decoding performance.

To further elucidate the relative contribution of specific bands, we trained LR models using features extracted exclusively from individual canonical bands, as opposed to using all canonical bands simultaneously. This approach allowed us to quantify the impact of each frequency band on decoding performance across the cohort, benchmarked against an all-band model.

Figure 10 presents the differences in AUC scores between models trained on individual bands and the model with access to all bands. Across all frequency bands, relying on only a single band resulted in an average AUC decrease of 18.8% ± 8.5% (mean ± standard deviation) compared to the all-band model. The smallest performance loss was observed in the low beta band (13–20 Hz), with an average AUC decrease of 15.25% ± 9.9%. The largest loss occurred in the high-frequency oscillation (HFO) band (200–500 Hz), with an average decrease of 23.01% ± 6.22%.

**Figure 10:**
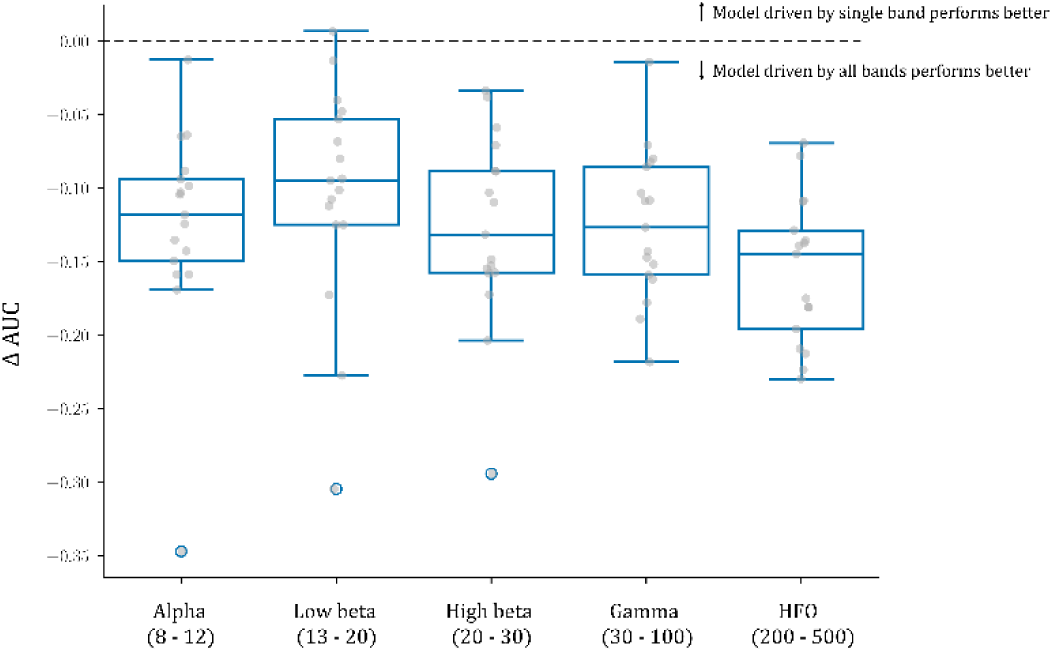
Impact of using individual frequency bands on decoding performance. Distribution plots show the loss in decoding performance (AUC difference) when models are trained using features from a single canonical frequency band compared to a model using all bands. Negative values indicate lower AUC for single-band models relative to the all-band model.

To further investigate whether contributions from different frequency bands are independent, we conducted an analysis examining the relationship between the decoding performance of models trained on single frequency bands and that of the all-band model. Specifically, we aimed to determine whether the AUC values from frequency-specific models could predict the AUC values of the all-band model, thereby assessing the extent to which individual bands contribute uniquely to the overall decoding performance.

We performed linear regression analyses where the AUC values obtained from models trained on individual frequency bands served as independent variables, and the AUC values from the all-band model were the dependent variable. By examining the strength and significance of these relationships, we could infer whether certain frequency bands had unique contributions to the decoding task that were not captured by other bands.

Our analyses revealed that the AUC values from individual-band models were indeed linearly related to those of the all-band model, explaining a statistically significant portion of the variance (*p* < 0.05). For instance, as shown in Figure 11, the AUC values from the low beta band model (13–20 Hz) explained 20% of the variance in the AUC values of the all-band model (R² = 0.20). Similarly, the gamma band model (30–100 Hz) explained 17% of the variance (R² = 0.17). These significant correlations indicate that both the low beta and gamma bands contribute meaningfully to the decoding performance achieved by the all-band model.

**Figure 11:**
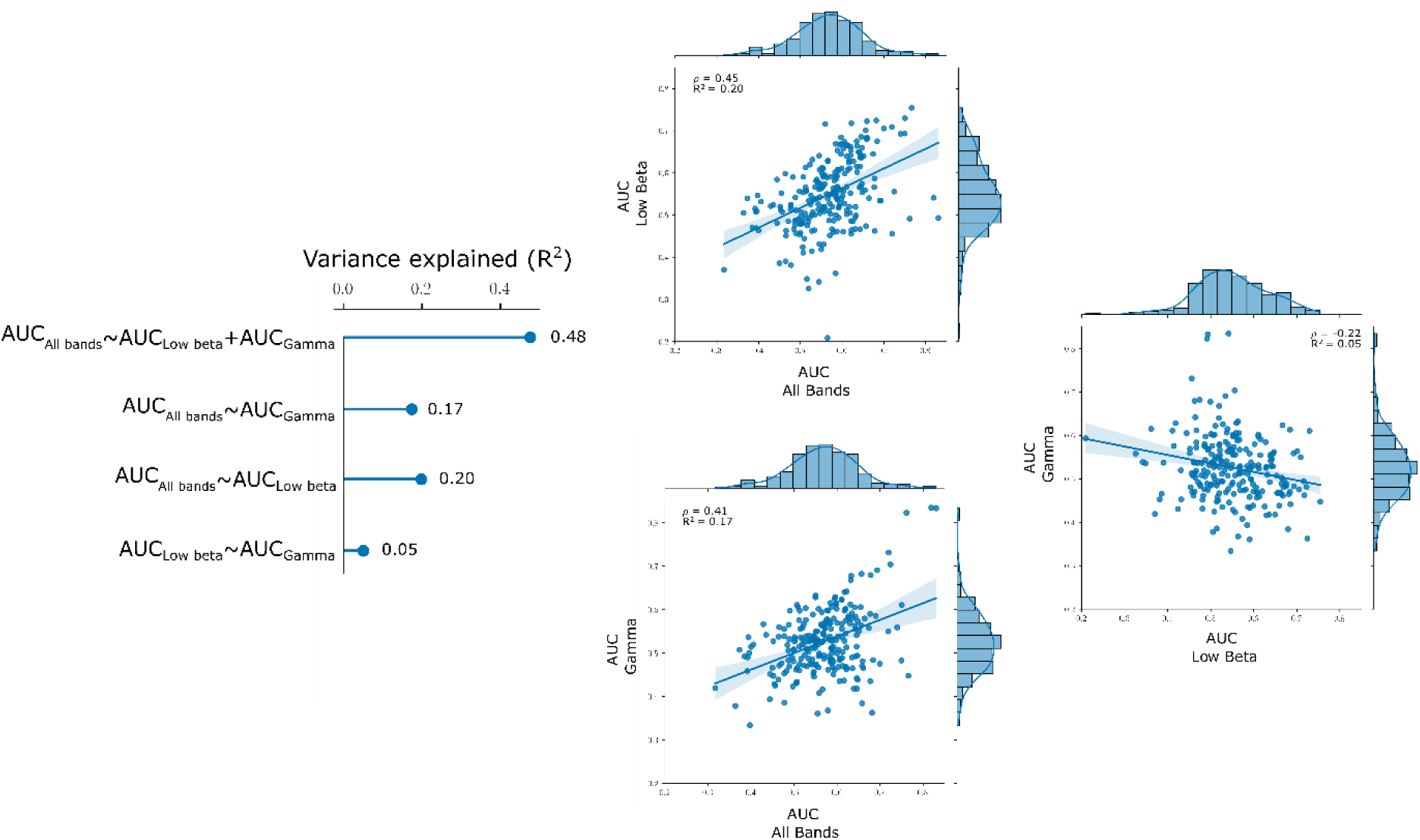
Scatter plots and linear regression lines showing the relationships between decoding performance (AUC values) of models trained on individual frequency bands and the all-band model. (top) All-band model vs. low beta band model. (bottom) All-band model vs. gamma band model. (right) Low beta band model vs. gamma band model.

Importantly, we observed that the AUC values from the low beta and gamma band models were poorly correlated with each other (R² = 0.05), suggesting that these frequency bands capture different aspects of the neural signals relevant to the decoding task. This low correlation implies that the contributions from the low beta and gamma bands are relatively independent of one another.

To assess the combined effect of these bands, we constructed a multiple regression model using the AUC values from both the low beta and gamma band models as independent variables to predict the AUC of the all-band model. This combined model explained 48% of the variance in the all-band model’s AUC (R² = 0.48), a substantial increase when compared to the variance explained by each band individually. The significant increase in explained variance when both bands are considered together indicates that the low beta and gamma bands provide complementary information that, when combined, enhances decoding performance.

These findings suggest that contributions from different frequency bands are both individually significant as well as relatively independent. The independence of these contributions implies that different frequency bands capture distinct neural dynamics associated with pre-movement periods. By leveraging the complementary information from multiple frequency bands, models can achieve superior decoding performance compared to using any single band alone. Our results highlight the importance of incorporating a broad spectrum of frequency bands in decoding models to capture the complex neural signatures underlying motor preparation and execution. The independent and additive contributions from different bands support the notion that neural processes involved in movement are distributed across multiple oscillatory activities.

Our findings align with previous literature highlighting the importance of multi-band analyses in decoding neural signals. In (Ahn et al., 2020), authors used microelectrodes recordings from the subthalamic nucleus of PD patients to decode a proxy for motor performance. They reported superior decoding performance when models were trained on a broad spectrum of frequencies compared to individual bands, consistent with our observations. Similarly, (Khawaldeh et al., 2020) and (He et al., 2021) emphasized the simultaneous contributions of multiple frequency bands in subthalamic and thalamic LFPs, as evidenced by the weights assigned by machine learning algorithms to features from various bands. (Khawaldeh et al., 2022) further demonstrated that including a broad range of frequency bands significantly improved the prediction of clinical movement impairment scores in Parkinson’s disease patients, compared to using only the canonical beta band.

For the specific task studied here—the detection of periods leading up to movement execution—we find that utilizing multiple frequency bands enhances decoding performance. These results support the multi-band, machine learning-based approach to neural decoding that has been proposed in the literature (Merk et al., 2022; Neumann & Rodriguez-Oroz, 2021; Neumann et al., 2019).

Our study also underscores the potential limitations of focusing solely on canonical frequency bands or predefined biomarkers. The variability in modulatory patterns across patients suggests that individualized models, which account for patient-specific neural dynamics across multiple frequency bands, may offer better performance. This highlights the importance of data-driven approaches in developing adaptive deep brain stimulation systems and other neurotechnological interventions.

### 4.2. Comparing Manually Extracted Features and Deep Learning-Based Features

One of the advantages of using parameterized deep learning systems, such as CNNs, is their ability to bypass the manual feature extraction process. While approaches based on manually extracted features require careful selection of features—such as choosing specific frequency bands for spectral analysis—the optimal set of features may vary between patients, making it challenging to generalize and potentially limiting performance.

Deep learning approaches accept raw data inputs and process them through a series of learned mathematical operations, in our case, convolutions applied to time-domain signals. These operations are parameterized and can capture a broad spectrum of signal characteristics. The parameters are optimized during the training process to minimize a loss function related to the classification task. This data-driven learning of feature representations eliminates the need for manual implementation of signal processing pipelines to reduce high-dimensional time-series data to meaningful feature sets.

In this study, we investigated whether deep learning systems could learn feature representations that capture the information necessary to decode pre-movement periods, and whether these learned features provide any advantages over manually extracted features. To this end, we implemented two deep learning-based systems: a standard CNN and a hybrid model referred to as FeatCNN. The CNN processes signals through a cascade of convolutional filters whose coefficients are learned during training, as described in Section 2.4. The FeatCNN incorporates a CNN as a subcomponent but additionally uses manually extracted features, thereby combining the two feature sets to make predictions.

By comparing the performance of these two systems, we aimed to determine whether manually extracted features contain relevant information that CNNs cannot capture, and conversely, whether CNNs can capture all pertinent information without the need for manual features. Figure 12 presents the paired comparison between the AUC values of the CNN and FeatCNN models. The results show no significant difference in decoding performance between the two models, suggesting that the addition of manually extracted features does not provide a substantial benefit over the CNN’s learned features (*p* > 0.05).

**Figure 12:**
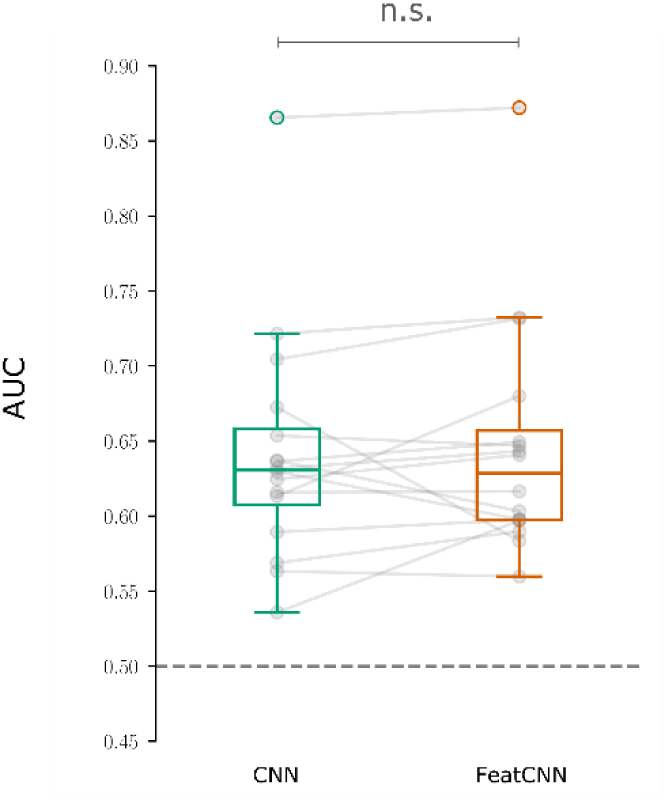
Paired comparison of AUC values for CNN and FeatCNN models across patients. Each pair of bars represents an individual patient. No significant difference in decoding performance was observed, indicating that the addition of manually extracted features does not enhance the CNN’s ability to decode pre-movement periods.

The lack of significant difference in performance between the CNN and FeatCNN indicates that the CNN is capable of capturing the relevant information content from the time-series signals through its learned convolutional filters, including modulatory patterns in relevant frequency bands or their correlates. This finding suggests that CNNs can effectively learn feature representations that are as informative as manually extracted features commonly used in adaptive DBS applications.

However, this does not imply that deep learning-based systems and systems based on manually extracted features are equivalent in how they process data or make inferences. To explore this further, we analysed the pairwise correlations between the performance metrics (AUC values) of the four models: LR, GBDT, CNN, and FeatCNN. Our analysis revealed stronger correlations within the deep learning-based systems (CNN and FeatCNN) and within the feature-based systems (LR and GBDT) than across the two categories. Specifically, 54% of the variance was shared between the CNN and FeatCNN models, and 52% between the LR and GBDT models. In contrast, the CNN shared only 17% and 18% of the variance with the GBDT and LR models, respectively.

Moreover, the correlations between the FeatCNN and the feature-based models (LR and GBDT) were higher (40% and 33%, respectively) than those between the CNN and the feature-based models, suggesting that the inclusion of manually extracted features in the FeatCNN model makes it operate more similarly to the feature-based systems. Nonetheless, the FeatCNN’s coupling remains tighter with the CNN than with the LR and GBDT models. These relationships are visualized in Figure 13A, which depicts the pairwise correlations between the models, and Figure 13B, which presents a two-dimensional embedding where the distances between points represent the proportion of variance explained.

**Figure 13:**
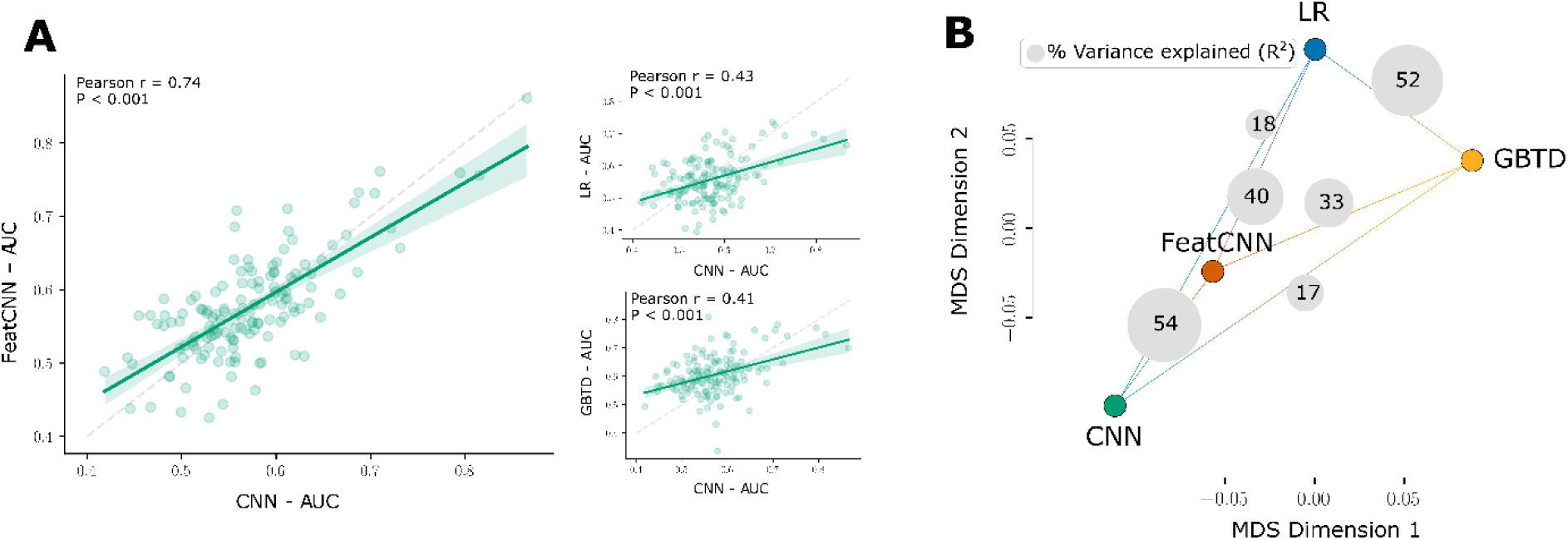
Similarity analysis of decoding performance across models. (A) Pairwise correlations between the AUC values of the models (higher correlation values indicate greater similarity in model performance). (B) Two-dimensional embedding representing the relationships between models based on the proportion of shared variance in AUC values. The distances between points reflect the degree of similarity in decoding performance across patients.

These findings suggest that, while the deep learning-based and feature-based models may achieve similar levels of decoding performance, they may rely on different aspects of the neural signals or process the information differently to make predictions. The deep learning models may capture complex, non-linear relationships and higher-order features that are not explicitly represented in the manually extracted feature sets. Conversely, feature-based models rely on predefined features that may be more interpretable but potentially less flexible in capturing individual patient variability.

The mechanisms underlying the differences between deep learning-based and feature-based models, as well as the potential implications for translational applications, remain areas for further research, and understanding these differences is crucial for developing reliable and interpretable models for clinical use.

### 4.3. Inference-Time Computational Cost

In the context of aDBS, signal processing pipelines that incorporate machine learning-based components present translationally exciting opportunities (Neumann et al., 2019). However, the computational environment within which aDBS can be feasibly implemented outside of the clinic remains constrained due to limitations in hardware resources and power consumption (Ansó et al., 2022). Therefore, efforts to develop algorithms intended for aDBS applications should take these considerations into account to ensure that the systems are practical for real-world deployment.

Estimating the computational complexity of algorithms implemented in high-level scripting languages such as Python or MATLAB is non-trivial, as these implementations may differ significantly from implementations optimised for the embedded systems that would be used in real-time applications. Nonetheless, providing reference values for the computational requirements can offer valuable insights into the feasibility of different algorithms in computationally constrained environments.

To assess the computational demands of our models, we benchmarked the time required to make a single inference-time prediction—that is, to process time-series signals and produce a model output. This benchmarking provides an approximate measure of the computational efficiency of each algorithm during real-time operation.

The feature extraction step, when using the full feature set, requires 187 milliseconds (ms) per prediction. Within this, the estimation of spectral power features—which drive most of the performance of the feature-based models (see Section 4.2) —takes only 5 ms. The time estimates for the forward pass of the different algorithms are outlined in Table 3.

**Table 3:**
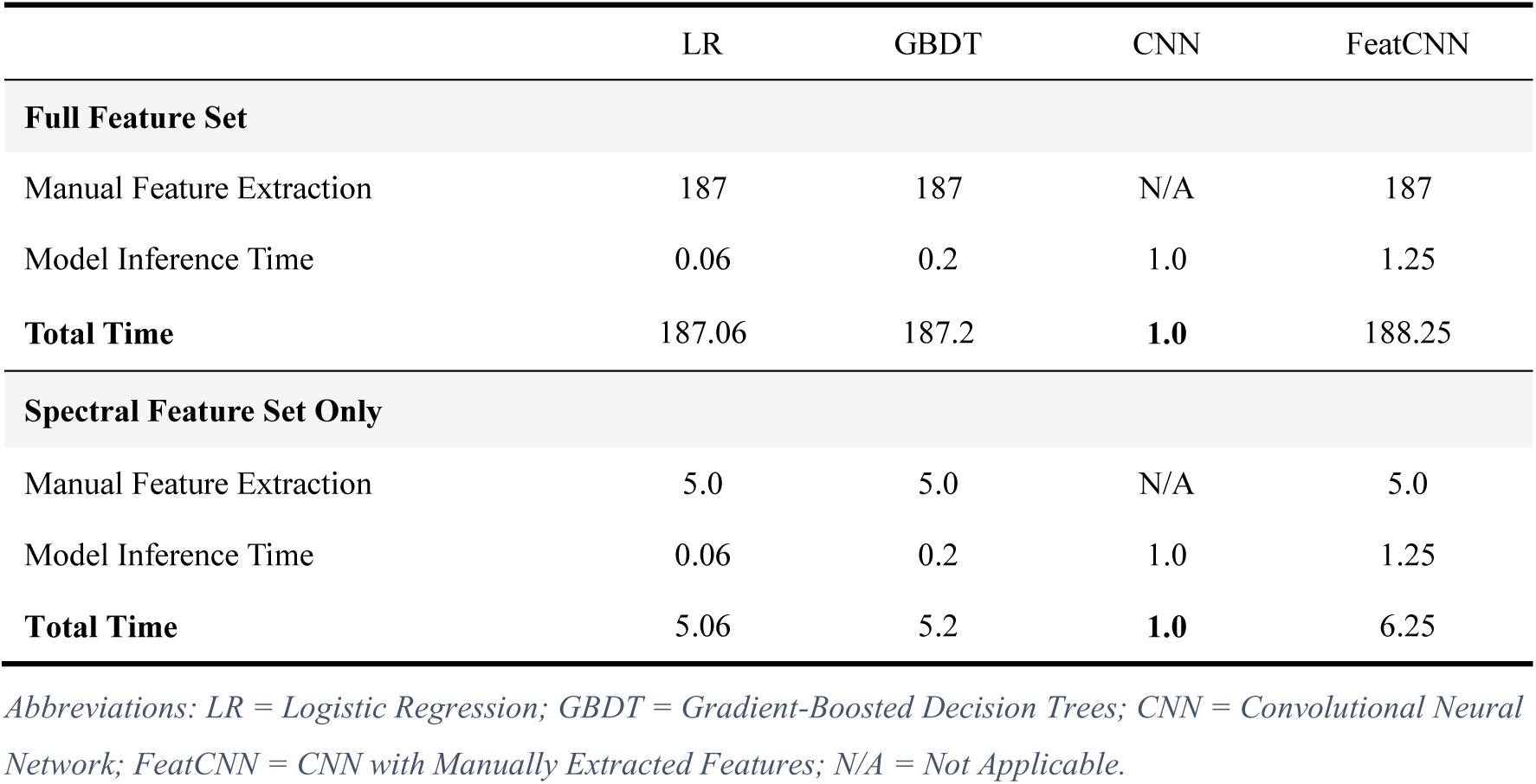
Benchmarked Time [in milliseconds] Required for an Inference-Time Prediction.

|  | LR | GBDT | CNN | FeatCNN |
| --- | --- | --- | --- | --- |
| **Full Feature Set** |  |  |  |  |
| Manual Feature Extraction | 187 | 187 | N/A | 187 |
| Model Inference Time | 0.06 | 0.2 | 1.0 | 1.25 |
| **Total Time** | **187.06** | **187.2** | **1.0** | **188.25** |
| **Spectral Feature Set Only** |  |  |  |  |
| Manual Feature Extraction | 5.0 | 5.0 | N/A | 5.0 |
| Model Inference Time | 0.06 | 0.2 | 1.0 | 1.25 |
| **Total Time** | **5.06** | **5.2** | **1.0** | **6.25** |

*Abbreviations: LR = Logistic Regression; GBDT = Gradient-Boosted Decision Trees; CNN = Convolutional Neural Network; FeatCNN = CNN with Manually Extracted Features; N/A = Not Applicable.*

When using the full feature set, which includes time-domain statistics, autoregressive coefficients, cepstral coefficients, and spectral features, the feature extraction step dominates the computational cost, requiring 187 ms per prediction. The LR and GBDT models have minimal inference times of 0.06 ms and 0.2 ms, respectively, resulting in total inference times of approximately 187 ms. In contrast, the CNN model, which operates directly on the raw time-series data without manual feature extraction, has a total inference time of 1.0 ms. The FeatCNN model, which combines the CNN with manually extracted features, requires both the feature extraction time (187 ms) and the CNN inference time (1.25 ms), leading to a total inference time of approximately 188 ms.

When using only the spectral feature set—which, as previously discussed, captures most of the relevant information—the feature extraction time is significantly reduced to 5 ms. Under this configuration, the total inference times for the LR and GBDT models decrease to approximately 5 ms, whereas the CNN’s inference time remains at 1.0 ms, as it does not rely on manual feature extraction. The FeatCNN model’s total inference time reduces to approximately 6.25 ms.

### 4.4. Limitations

In this study, we benchmarked the ability to decode upper limb movements in the milliseconds preceding movement execution by implementing and testing various machine learning algorithms and feature sets. While our findings provide valuable insights, several limitations should be acknowledged.

First, the sample size of our cohort was relatively small (n = 11), which may limit the generalizability of the results. The heterogeneity in modulatory patterns observed across patients led to variability in decoding performance, particularly in cross-patient models. This inter-patient variability is further highlighted by the poor performance of models trained across patients compared to individualized, patient-specific decoders. Including a larger and more diverse patient population in future studies would increase the statistical power and help identify robust, cross-patient patterns that could enhance model generalizability.

Second, we employed limited machine learning interpretability techniques on our models—which included logistic regression, gradient-boosted decision trees, and convolutional neural networks. The lack of significant correlations between modulation indices in specific frequency bands and decoding performance suggests that complex, non-linear relationships may exist in the data that are not fully understood. Employing explainable machine learning approaches or incorporating interpretability methods could provide deeper insights into the neural mechanisms underlying movement preparation and execution.

Third, all analyses were performed offline, despite being implemented with a real-time compatible and fully causal pipeline. This means that we lack performance metrics gathered during real-time aDBS. Assessing the models in real-time settings is crucial to evaluate their practical utility and effectiveness in clinical environments. Future work should focus on real-time implementation and testing to determine the feasibility and reliability of these decoding systems in actual aDBS applications.

Additionally, while we investigated the use of both thalamic LFPs and scalp EEG signals, we did not extensively explore the potential differences and complementarities between these modalities. Further research could delve into how combining LFP and EEG data might enhance decoding performance or provide more robust biomarkers for movement prediction.

Finally, the experimental tasks used to elicit movements were specific and may not encompass the full range of motor activities encountered in daily life. This could limit the applicability of our models to more naturalistic settings. Expanding the repertoire of tasks and incorporating more ecologically valid movement paradigms could improve the models’ relevance and utility in real-world scenarios.

### 4.5. Conclusion

We demonstrated that early neural correlates of upper limb movement within thalamic LFP and EEG signals can be leveraged to decode pre-movement periods prior to movement execution. Decoding performance was statistically significant from chance at approximately 680 ms before movement onset for LFPs and 1.09 s for EEGs. Individualized, patient-specific decoders outperformed cross-patient models, consistent with the observed variability in modulatory patterns across subjects. Multiple frequency bands contributed independently to decoding performance, highlighting the importance of incorporating information from a broad range of frequencies.

To optimize decoding performance, our findings emphasize the necessity of individualized approaches that account for patient-specific neural dynamics and leverage multi-band spectral information. These insights have important implications for the development of adaptive neurostimulation therapies and brain-machine interfaces, suggesting that personalized, data-driven models can enhance the effectiveness and reliability of neural decoding systems in clinical applications.

## Data Availability

All data produced in the present study are available upon reasonable request to the authors.

## Funding

This work was supported by the Medical Research Council (MC_UU_0003/2, MR/V00655X/1, MR/P012272/1), the Medical and Life Sciences Translational Fund (MLSTF) from the University of Oxford, the National Institute for Health Research (NIHR) Oxford Biomedical Research Centre (BRC), and the Rosetrees Trust, UK. S.H. was supported by the Guarantors of Brain and the Royal Society Sino-British Fellowship Trust (IES\R3\213123).

R.S.S was personally supported by an NIHR CL award (CL-2021-16-1502) and an RCS England Pump Priming grant funded by Saven Research & Development Fund.

## Competing interests

The authors declare no competing interests.

## Notes

### Competing Interest Statement

The authors have declared no competing interest.

### Author Declarations

Ethics committee of South Central-Oxford B Research Ethics Committee gave ethical approval for this work.

